# The potential impact of reductions in international donor funding on tuberculosis in low- and middle-income countries

**DOI:** 10.1101/2025.04.23.25326313

**Authors:** Rebecca A. Clark, C. Finn McQuaid, Alexandra S. Richards, Roel Bakker, Tom Sumner, Tomos Prŷs-Jones, Rein M. G. J. Houben, Richard G. White, Katherine C. Horton

**Affiliations:** Department of Infectious Disease Epidemiology, London School of Hygiene and Tropical Medicine, London, UK; TB Modelling Group, London School of Hygiene and Tropical Medicine, London, UK

## Abstract

**Background:** Tuberculosis services in many settings rely heavily on international donor funding. In 2025, the United States Agency for International Development (USAID) was dismantled, and other countries also announced cuts to overseas development assistance. We quantified potential epidemiological impacts attributable to these reductions in international donor funding.

**Methods:** We calibrated a deterministic tuberculosis model to epidemiological indicators in low- and middle-income countries. We projected three future scenarios assuming: a) levels of funding in 2024 continue through 2035, b) termination of USAID funding from 2025, and c) additional reductions in funding through The Global Fund in line with current donor announcements from 2025. We assumed a reduction in tuberculosis treatment initiation rates proportional to budget reductions for each scenario, estimating cumulative excess incident episodes of symptomatic tuberculosis and tuberculosis deaths.

**Findings:** We modelled 79 countries, representing 91% of global tuberculosis incidence and 90% of global tuberculosis mortality in 2023. Our modelling suggested that the termination of USAID funding may lead to 420 500 excess tuberculosis deaths by 2035. Further reductions in funding in line with current announcements by the United States, France, the United Kingdom, and Germany may lead to an additional 699 200, 63 100, 50 500, and 30 500 TB deaths, respectively. Impacts would be greatest in low-income countries.

**Interpretation:** We estimate substantial potential impacts on tuberculosis morbidity and mortality due to reductions in international donor funding. Expanded support from domestic and international donors is essential to address immediate gaps in prevention, diagnosis, and treatment.

**Funding:** This work was unfunded.

## Introduction

An estimated 10.8 million people developed tuberculosis (TB) in 2023, and 1.25 million affected individuals died in the same year.^1^ Although TB remains the leading infectious cause of mortality globally, tremendous progress has been made to reduce morbidity and mortality. Between 2010 and 2020, estimated incidence fell by 20% and mortality by 40%.^1^ This progress was interrupted by the COVID-19 pandemic, but recovery efforts have been strong, and the treatment coverage gap is now the smallest it has been since reporting began.^1^ However, reaching the ambitious EndTB targets of a 90% decline in incidence and a 95% decline in mortality by 2035^2^ would require large shifts in current trends, for which funding remains a major challenge.^1^

TB disproportionately affects low- and middle-income countries,^3^ and prevention, diagnostic, and treatment services in many settings rely heavily on international donor funding. The largest bilateral donor to the TB response has been the United States Agency for International Development (USAID), which contributed 19% of international donor funding reported by national TB programmes (NTPs) in 2022. However, The Global Fund to Fight AIDS, TB and Malaria (The Global Fund) has been the primary source of international donor funding for TB since 2013, providing 66% of international donor funding reported by NTPs in 2022.^1^ While more than 80 countries contribute to The Global Fund, the United States has been the largest donor. In the Seventh Replenishment (2023–2025), the US pledged 38% of total pledges, followed by France (10%), Germany (8%), and the United Kingdom (7%).^4^ Historic contributions to TB prevention and care by these agencies have saved the lives of millions people with TB.^5^

In the first quarter of 2025, the United States abruptly dismantled USAID and cancelled funding for bilateral aid through the agency.^6^ Around the same time, other countries also announced reductions in contributions to bilateral aid programmes and multinational organisations. The United Kingdom, France, the Netherlands, and Belgium have announced cuts to overseas development assistance budgets ranging from 25% (in Belgium) to 40% (in the UK).^7^

The impact of USAID funding cuts has already interrupted TB prevention, diagnostic, and treatment services,^8,9^ but the long-term impacts of these disruptions has not been evaluated. We used mathematical modelling to estimate the epidemiological impacts attributable to the termination of USAID funding and the potential impact of future cuts in donor funding through The Global Fund in low- and middle-income countries.

## Methods

We used a dynamic, compartmental model of *Mycobacterium tuberculosis* (*Mtb*) transmission, progression, and care to simulate TB epidemic trajectories in selected low- and middle-income countries (Supplemental Figure 1).^10^

### Calibration

For each country, we calibrated the model to nine epidemiological targets in 2023: the country-specific TB incidence rate (for all ages, those aged 0–14 years, and those 15 years and older, separately), country-specific TB notification rate (for all ages, those aged 0–14 years, and those 15 years and older, separately), country-specific TB mortality rate (for all ages), and the global fraction of asymptomatic TB among infectious TB (asymptomatic + symptomatic). Models for countries classified as having a high TB burden due to HIV were fit to at least three additional country-specific all-age targets in 2019: HIV prevalence, anti-retroviral therapy (ART) coverage, TB incidence rate in people living with HIV, and TB mortality rate in people living with HIV.

Calibration was conducted using history matching with emulation using the hmer package in R,^11^ generating 200 fitted parameter sets per country. We used the distribution of results produced by these parameter sets to quantify parameter uncertainty.^12^

### Scenarios

For each country, we projected three future scenarios. We first simulated a business-as-usual scenario assuming funding and programmatic activities continue at 2024 levels. Second, we then modelled a scenario representing the impact of the historic termination of USAID funding to NTPs from 2025.

Third, we simulated an additional reduction in funding through The Global Fund from 2025. For each donor that pledged at least 1% of total pledged donations for The Global Fund’s Seventh Replenishment (2023–2025),^4^ we estimated the impact of changes funding in line with announced changes in overseas development assistance.^13-17^ When considering the total impact of expected reductions in overseas development assistance, we excluded countries expected to increase their contributions. We also estimated the impact of complete termination of funding to The Global Fund by donor. We assumed all funding cuts were introduced in 2025, sustained into the future, and not replaced by other funding sources.

Funding cuts are likely to have a direct impact on TB treatment by limiting the accessibility of TB services and the availability of TB diagnostics and treatment. Thus, we assumed a reduction in treatment initiation rate proportional to budget reduction in each scenario. Budget reductions were calculated using budget data from the World Health Organization (WHO), which has monitored funding for TB programmes since 2022.^1^ For each country, WHO data were used to calculate the proportion of total expected funding for all budget line items from USAID and from The Global Fund in 2023. We disaggregated The Global Fund contributions by donor using data on pledged donations for The Global Fund’s Seventh Replenishment (2023-2025).^4^

### Epidemiologic impact

For each scenario, we calculated the cumulative number of symptomatic TB episodes and TB deaths between 2025 and 2035 for each scenario compared to business-as-usual. Results are presented as median values and 95% uncertainty intervals (UIs). We evaluated outcomes across all countries included and grouped results by WHO region and World Bank income group.

### Data accessibility

Replication data and analysis scripts are available on GitHub at https://github.com/lshtm-tbmg/tbmod-pub-funding-impacts.

## Findings

### Model calibration

Of 111 low- and middle-income countries with data available to attempt calibration, complete budget data were available for 90 countries.^18^ Model calibrations were completed for 79 of 90 countries (Figure 1), representing 91% of global TB incidence and 90% of global TB mortality in 2023.^1^ Of these, 43 countries received bilateral aid from USAID, 23 received donor funding from both USAID and The Global Fund in 2023, and 17 did not receive funding from either source.^18^ No countries received funding from The Global Fund without also receiving funding from USAID.

**Figure 1:**
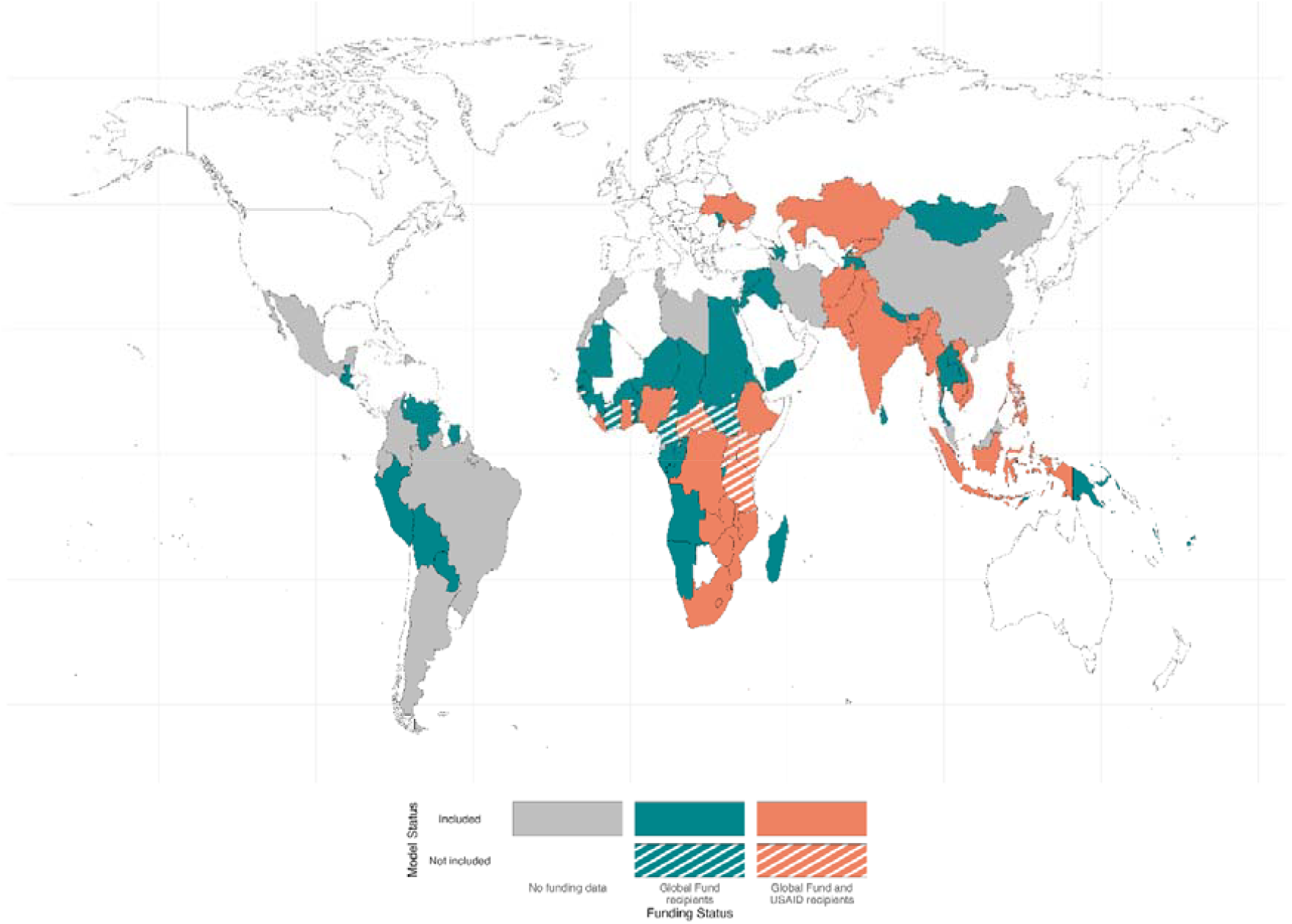
World map showing countries that were modelled and received funding from the United States Agency for International Development (USAID) and The Global Fund to Fight AIDS, Tuberculosis and Malaria (The Global Fund) (solid orange), The Global Fund (solid green), or received no funding from either source (grey). Countries that were not modelled but received funding from USAID and The Global Fund (striped orange) or The Global Fund (striped green) are also shown.

Of the 79 countries included in the analysis, the highest number of countries were from the WHO African region (n=29), followed by the WHO Region of the Americas (n=13) and the WHO Eastern Mediterranean Region (n=12) (Supplemental Table 1). According to World Bank income groups, 34 countries were classified as lower-middle income countries, 29 were classified as upper-middle income countries, and the remaining 16 as low-income countries.

### Impact of termination of USAID funding

We estimated that termination of funding from USAID to NTPs may result in 1.4 million (UI 1.1, 1.7) excess episodes of symptomatic TB and 420 500 (UI 335 800, 558 000) excess TB deaths by 2035, a 1·3% and 3·8% increase respectively over the business-as-usual scenario assuming funding and programmatic activities continue at 2024 levels (Figure 2, Table 1). This reversal in trends would erase twelve years of progress in reducing TB incidence and a decade of progress in reducing TB mortality by 2035.

**Table 1:** Excess symptomatic tuberculosis (TB) episodes and TB deaths (2025–2035) projected to result from termination in funding to national TB programmes from the United States Agency for International Development (USAID) and reductions in funding from The Global Fund to Fight AIDS, Tuberculosis and Malaria (The Global Fund) by donors that contribute at least 1% of total pledges

| Donor | Change in funding | Excess symptomatic tuberculosis episodes |  | Excess tuberculosis deaths |  |
| --- | --- | --- | --- | --- | --- |
|  |  | median (95% UI) | % (95% UI) | median (95% UI) | % (95% UI) |
| Termination of funding via USAID |  |  |  |  |  |
| United States | -100% | 1 390 900<br>(1 113 500, 1 675 000) | 1.3%<br>(1.0, 1.7) | 420 500<br>(335 800, 558 000) | 3.8%<br>(3.0, 4.9) |
| Expected change in funding via The Global Fund |  |  |  |  |  |
| Total reductions | .. | 3 928 100<br>(2 935 400, 5 050 700) | 3.6%<br>(2.6, 5.0) | 970 300<br>(698 100, 1 217 800) | 8.4%<br>(6.2, 10.8) |
| United States | -93% | 2 841 700<br>(2 135 800, 3 652 700) | 2.6%<br>(1.9, 3.6) | 699 200<br>(505 600, 869 200) | 6.0%<br>(4.5, 7.7) |
| France | -37% | 260 200<br>(196 500, 329 100) | 0.2%<br>(0.2, 0.3) | 63 100<br>(46 200, 77 900) | 0.6%<br>(0.4, 0.7) |
| Germany | -24% | 125 800<br>(95 000, 159 100) | 0.1%<br>(0.1, 0.2) | 30 500<br>(22 400, 37 700) | 0.3%<br>(0.2, 0.3) |
| United Kingdom | -40% | 207 900<br>(157 000, 263 000) | 0.2%<br>(0.1, 0.3) | 50 500<br>(36 900, 62 300) | 0.4%<br>(0.3, 0.6) |
| Japan | +17% | -79 700<br>(-100 700, -60 200) | -0.1%<br>(-0.1, -0.1) | -19 400<br>(-23 900, -14 200) | -0.2%<br>(-0.2, -0.1) |
| Canada | -26% | 103 300<br>(78 000, 130 600) | 0.1%<br>(0.1, 0.1) | 25 100<br>(18 400, 30 900) | 0.2%<br>(0.2, 0.3) |
| European Commission | -35% | 109 200<br>(82 500, 138 100) | 0.1%<br>(0.1, 0.1) | 26 500<br>(19 400, 32 700) | 0.2%<br>(0.2, 0.3) |
| Sweden | -10% | 12 000<br>(9 000, 15 100) | 0.0%<br>(0.0, 0.0) | 2 900<br>(2 100, 3 600) | 0.0%<br>(0.0, 0.0) |
| Norway | -12% | 10 100<br>(7 600, 12 800) | 0.0%<br>(0.0, 0.0) | 2 500<br>(1 800, 3 000) | 0.0%<br>(0.0, 0.0) |
| Italy | +15% | -12 000<br>(-15 200, -9 100) | 0.0%<br>(0.0, 0.0) | -2 900<br>(-3 600, -2 100) | 0.0%<br>(0.0, 0.0) |
| Netherlands | -29% | 22 700<br>(17 100, 28 600) | 0.0%<br>(0.0, 0.0) | 5 500<br>(4 000, 6 800) | 0.0%<br>(0.0, 0.1) |
| Australia | -3% | 2 300<br>(1 800, 2 900) | 0.0%<br>(0.0, 0.0) | 600<br>(400, 700) | 0.0%<br>(0.0, 0.0) |

**Figure 2:**
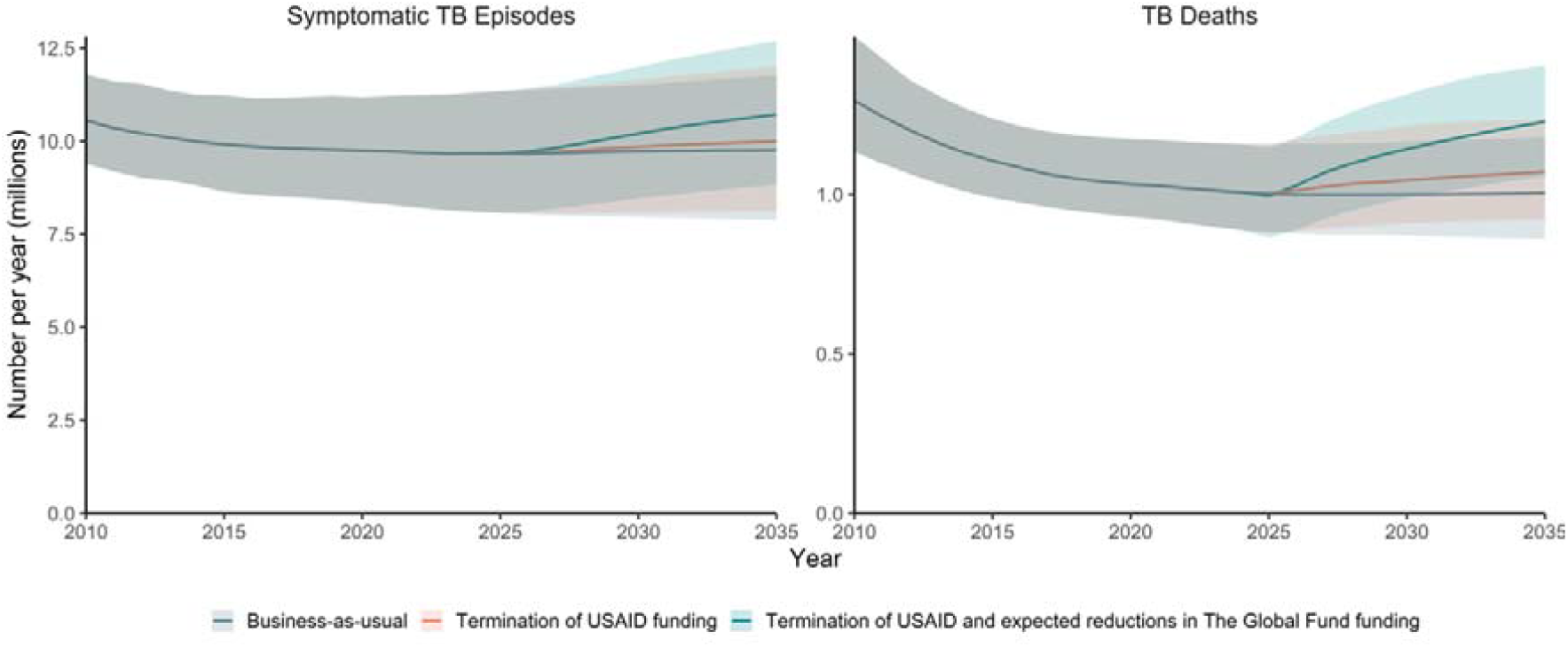
Excess symptomatic tuberculosis (TB) episodes and TB deaths (2025–2035) projected to result from termination in funding to national TB programmes from the United States Agency for International Development (USAID) and reductions in funding from The Global Fund to Fight AIDS, Tuberculosis and Malaria (The Global Fund) by donors that contribute at least 1% of total pledges

The absolute and relative impact was greatest in the WHO African region, with an estimated 748 100 (UI 553 300, 952 000) excess episodes of symptomatic TB by 2035 (3·3% increase compared to business-as-usual) and 286 800 (UI 215 700, 394 200) excess TB-associated deaths (10·2% increase compared to business-as-usual) (Table 2). The next greatest impact was in the WHO South-East Asia Region, followed by the Western Pacific Region, the Eastern Mediterranean Region, and the European Region. The WHO Region of the Americas did not receive funding from USAID.

**Table 2:** Excess symptomatic tuberculosis (TB) episodes and TB deaths (2025–2035) projected to result from termination in funding to national TB programmes from the United States Agency for International Development (USAID) and reductions in funding from The Global Fund to Fight AIDS, Tuberculosis and Malaria (The Global Fund) by donors that contribute at least 1% of total pledges by World Health Organization region and World Bank income group

|  |  | Excess symptomatic tuberculosis episodes |  | Excess tuberculosis deaths |  |
| --- | --- | --- | --- | --- | --- |
|  |  | median (95% UI) | % (95% UI) | median (95% UI) | % (95% UI) |
| **Termination of funding via USAID** |  |  |  |  |  |
| **World Health Organization Region** | **African Region** | 748 100<br>(553 300, 952 000) | 3.3%<br>(2.1, 4.6) | 286 800<br>(215 700, 394 200) | 10.2%<br>(7.5, 12.9) |
|  | **Region of the Americas** | .. | .. | .. | .. |
|  | **Eastern Mediterranean Region** | 32 400<br>(17 100, 48 100) | 0.4%<br>(0.2, 0.6) | 9 200<br>(5 600, 14 800) | 1.2%<br>(0.7, 1.9) |
|  | **European Region** | 4 200<br>(2 100, 6 800) | 0.7%<br>(0.3, 1.1) | 200<br>(-1 800, 1 500) | 0.2%<br>(-2.1, 1.6) |
|  | **South-East Asia Region** | 437 400<br>(300 200, 615 600) | 0.8%<br>(0.6, 1.2) | 109 100<br>(53 400, 151 400) | 1.8%<br>(0.9, 2.6) |
|  | **Western Pacific Region** | 165 000<br>(80 200, 262 100) | 0.9%<br>(0.4, 1.5) | 15 400<br>(-6 400, 34 700) | 1.6%<br>(-0.6, 3.4) |
| **World Bank Income Group** | **Low-income** | 529 700<br>(359 000, 698 500) | 4.6%<br>(2.6, 7.0) | 200 500<br>(148 300, 289 800) | 15.5%<br>(11, 19.9) |
|  | **Lower-middle income** | 639 200<br>(492 300, 790 700) | 1.0%<br>(0.7, 1.3) | 186 400<br>(124 600, 263 400) | 2.7%<br>(1.8, 3.7) |
|  | **Upper-middle income** | 220 100<br>(100 800, 347 000) | 0.8%<br>(0.4, 1.3) | 30 900<br>(-6 300, 55 900) | 1.1%<br>(-0.2, 2.0) |
| **Expected reductions in funding via The Global Fund** |  |  |  |  |  |
| **World Health Organization Region** | **African Region** | 1 016 200<br>(741 100, 1 291 100) | 4.3<br>(2.8, 6.3) | 391 500<br>(279 700, 529 600) | 12.5<br>(8.4, 15.9) |
|  | **Region of the Americas** | 8 500<br>(5 500, 10 900) | 0.3<br>(0.2, 0.4) | 300<br>(-1 200, 1 900) | 0.1<br>(-0.4, 0.6) |
|  | **Eastern Mediterranean Region** | 805 300<br>(450 000, 1 228 200) | 8.6<br>(4.4, 13.9) | 196 300<br>(132 100, 289 900) | 25.9<br>(16.2, 35.3) |
|  | **European Region** | 9 800<br>(5 100, 15 400) | 1.5<br>(0.6, 2.3) | 800<br>(-3 500, 4 000) | 0.8<br>(-4.5, 3.9) |
|  | **South-East Asia Region** | 1 807 200<br>(974 000, 2 649 400) | 3.4<br>(1.9, 5.4) | 348 600<br>(87 300, 539 500) | 5.6<br>(1.4, 9.1) |
|  | **Western Pacific Region** | 291 100<br>(177 100, 421 400) | 1.5<br>(0.9, 2.4) | 38 700<br>(2 000, 72 100) | 4.1<br>(0.2, 7.5) |
| **World Bank Income Group** | **Low-income** | 731 800<br>(521 300, 964 500) | 6.2<br>(3.9, 9.0) | 268 900<br>(196 800, 389 100) | 18.1<br>(12.2, 22.9) |
|  | **Lower-middle income** | 1 902 600<br>(1 429 700, 2 362 800) | 2.9<br>(1.9, 3.9) | 542 200<br>(392 500, 708 400) | 7.5<br>(5.5, 10.3) |
|  | **Upper-middle income** | 1 298 500<br>(506 800, 2 132 400) | 4.5<br>(1.8, 7.8) | 161 200<br>(-67 500, 333 600) | 5.9<br>(-2.6, 12.1) |

The absolute impact on episodes of symptomatic TB was greatest in lower-middle income countries, with an estimated 639 200 (UI 492 300, 790 700) excess episodes predicted between 2025–2035 compared to business-as-usual. However, the greatest relative impact on symptomatic TB episodes was in low-income countries (4·6% increase compared to business-as-usual). The greatest absolute and relative impact on excess TB-associated deaths was also in low-income countries, predicting an estimated 200 500 (UI 148 300, 289 800) excess TB deaths (15·5% increase compared to business-as-usual) (Table 2).

### Impact of potential future reductions in The Global Fund funding

We estimated that on top of termination of funding from USAID to NTPs, reductions in funding through The Global Fund could drastically increase the number of symptomatic TB episodes and TB-associated deaths by 2035. Reductions in funding in line with current announcements by the United States, France, the United Kingdom, and Germany may lead to an additional 2·8 million, 260 200, 207 900, and 125 800 episodes of symptomatic TB and an additional 699 200, 63 100, 50 500, and 30 500 TB deaths, respectively (Table 1). Expected reductions in funding across all donors who pledge at least 1% of total pledges to The Global Fund may result in an additional 3·9 million (UI 2·9, 5·1) episodes of symptomatic TB and 1·0 million (UI 0·7, 1·2) TB deaths, a 3·6% and 8·4% increase respectively over termination of USAID funding. This reversal in trends would erase more than 15 years of progress in reducing TB incidence and 13 years of progress in reducing TB mortality by 2035.

Total termination of all funding through The Global Fund to NTPs may result in a further 11·8 million (UI 8·7, 15·1) excess episodes of symptomatic TB and 3·0 million (UI 2·0, 4·0) excess TB deaths by 2035 (Supplemental Figure 2). These estimates are equivalent to increases of 10·8% and 26·1%, respectively, relative to projections from the scenario in which only funding from USAID is terminated. 3·0 million deaths may result from termination of funding to The Global Fund from the United States, 175 300 from France, 130 400 from Germany, 128 600 from the United Kingdom, and 117 700 from Japan (Supplemental Table 2).

## Discussion

The sudden termination of USAID funding risks reversing progress that has been made in recent years towards EndTB targets. Just as the burden of TB is inequitably distributed, we project the cessation of USAID funding will have the largest impact on low-income countries and countries in the African region, which have been most reliant on international donor funding. Potential further reductions in funding due to reductions in overseas development assistance, modelled here through potential cuts in contributions to The Global Fund, would likely further increase TB morbidity and mortality. These reductions would also disproportionately affect low-income countries and, in this analysis, countries in the Eastern Mediterranean region.

The impact of reductions international donor funding is inequitably distributed between countries, varying within income group and geographical region. We observe a relatively small average proportional increase in symptomatic TB and TB deaths attributable to termination of USAID funding modelled here, compared to the business-as-usual scenario assuming funding and programmatic activities continue at 2024 levels. This is likely because USAID contributions were low among countries with the estimated highest number of incident TB cases in 2023 (3% of total NTP budget in India, 6% in Indonesia, 0% in China, 13% in Philippines, and <1% in Pakistan). However, USAID contributions represented over a third of total NTP budgets in the Democratic Republic of Congo and Ethiopia. Donor funding through The Global Fund supports even greater proportions of NTP budgets in many countries, representing over 90% of NTP budgets in 11 countries modelled here, including Viet Nam, Madagascar, and South Sudan, and over half of NTP budgets in 29 countries modelled. Here, the impacts are larger.

The US has been a leader in global health investment for decades and, through USAID and The Global Fund, has contributed nearly half of all international donor funding to NTPs.^1^ These contributions have saved millions of lives.^5^ Our findings highlight the vulnerability of programmes that rely heavily on a single donor and support calls to diversify funding sources and reduce donor dependency.^19-21^ There is an urgent need to increase domestic funding for TB prevention, diagnostic, and treatment services to build more robust systems.^20,21^ However, support from other governments and philanthropic organisations must continue whilst these systems develop. The sudden departure of the US from the funding landscape requires expanded support from other donors to address immediate gaps in prevention, diagnosis, and treatment.

The upcoming Eighth Replenishment of The Global Fund will be crucial. If the announced overseas development assistance cuts are reflected in reduced contributions to The Global Fund, we estimate further morbidity and mortality beyond that attributed to USAID cuts. It is crucial that government donors fulfil their pledges to The Global Fund’s 2023-2025 funding cycle and uphold, or increase, their support in the Eighth Replenishment (2026-2028) meeting later this year.

Our analysis focuses only on direct international donor funding to NTPs, emphasising impacts on diagnosis and treatment without quantifying impacts on other aspects of TB prevention and care. We also have not modelled the impact of mitigation strategies that affected countries may implement in response to reductions in funding. We have not considered the impact of reductions in funding to HIV or ART programmes through USAID, the US President’s Emergency Plan for AIDS Relief (PEPFAR), or The Global Fund.^22,23^ Given HIV infection is the strongest risk factor for TB, and TB is the leading cause of death for people living with HIV, these funding cuts could have catastrophic effects, reversing the decades of declines in TB seen over the past decades, particularly for the African region. Additionally, we have not considered the impact of the elimination of funding to support health systems that might have indirect impacts on TB programmes, nor to multinational organisations that provide technical assistance and strategic support to affected countries. Conversely, we have only modelled scenarios in which funding reductions are sustained into the future; we have not considered alternative funding sources may mitigate impacts. Data were not available to quantify bilateral aid from countries other than the United States, so our analysis only considers contributions through The Global Fund for these countries. Our results rely on assumptions about the way changes in international donor funding impact TB prevention, diagnosis, and treatment and how this is implemented in the model; different assumptions lead to different projections.^24^

We estimate substantial impacts due to recent terminations and expected reductions in international donor funding that could reverse years of progress to reduce TB morbidity and mortality. Expanded support from domestic and international donors is essential to address immediate gaps in prevention, diagnosis, and treatment, and more robust funding systems are needed going forward.

## Funding statement

This work was unfunded. We used models that were developed with funding from the Wellcome Trust (310728/Z/24/Z, 218261/Z/19/Z), NIH (1R01AI147321-01, G-202303-69963, R-202309-71190), EDTCP (RIA208D-2505B), UK MRC (CCF17-7779 via SET Bloomsbury), ESRC (ES/P008011/1), BMGF (INV-004737, INV-035506), Open Philanthropy (GV673606227), and the WHO (2020/985800-0).

Authors are funded for other work as follows: RAC by BMGF (INV-001754) and NIH (G-202303-69963, R-202309-71190); CFM by BMGF (TB MAC OPP1135288, INV-059518), WHO (APW 203462345) and NIH (R-202309-71190); ASR by FCDO (Leaving no-one behind: transforming gendered pathways to health for TB); RMGJH by NIH (R-202309-71190, R01AI147321), NIHR (NIHR156644) and WT (310728/Z/24/Z); RGW by the Wellcome Trust (310728/Z/24/Z, 218261/Z/19/Z), NIH (1R01AI147321-01, G-202303-69963, R-202309-71190), EDTCP (RIA208D-2505B), UK MRC (CCF17-7779 via SET Bloomsbury), ESRC (ES/P008011/1), BMGF (INV-004737, INV-035506), Open Philanthropy (GV673606227), and the WHO (2020/985800-0); KCH by NIH (R-202309-71190) and FCDO (Leaving no-one behind: transforming gendered pathways to health for TB).

## Author contributions

RAC: Conceptualisation, Software, Formal analysis, Visualisation, Writing - Original Draft. CFM: Conceptualisation, Methodology, Writing - Review & Editing. ASR: Methodology, Visualisation, Writing - Review & Editing. RB: Software, Writing - Review & Editing. TS: Software, Writing - Review & Editing. TP: Software, Writing - Review & Editing. RMGJH: Conceptualisation, Writing - Review & Editing. RGW: Conceptualisation, Software, Writing - Review & Editing. KCH: Conceptualisation, Methodology, Writing - Original Draft.

## Competing interests

Authors have no competing interests.

