## Supplementary Material for "The potential impact of reductions in international donor funding on tuberculosis in low- and middle-income countries"

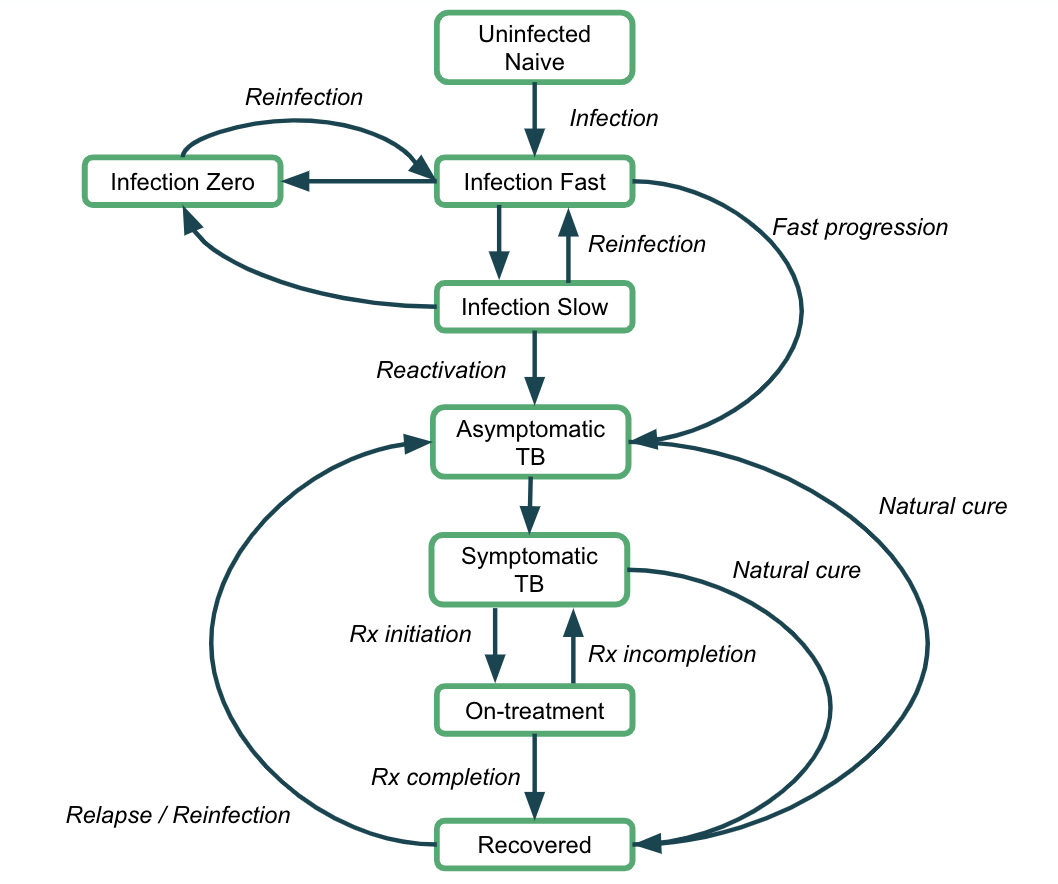

**Supplemental Figure 1**: Model structure

**Supplemental Table 1**: Number of countries modelled by and World Bank Income Group and World Health Organization Region (WHO).

|  | | **World Bank Income Groups** | | | **Total** |
| --- | --- | --- | --- | --- | --- |
|  |  | **Low-income** | **Lower-middle income** | **Upper-middle income** |  |
| **World Health Organization Region** | **African Region** | 12 | 13 | 4 | 29 |
|  | **Region of the Americas** | 0 | 1 | 12 | 13 |
|  | **Eastern Mediterranean Region** | 4 | 5 | 3 | 12 |
|  | **European Region** | 0 | 2 | 4 | 6 |
|  | **South-East Asia Region** | 0 | 7 | 2 | 9 |
|  | **Western Pacific Region** | 0 | 6 | 4 | 10 |
| **Total** | | 16 | 34 | 29 | 79 |

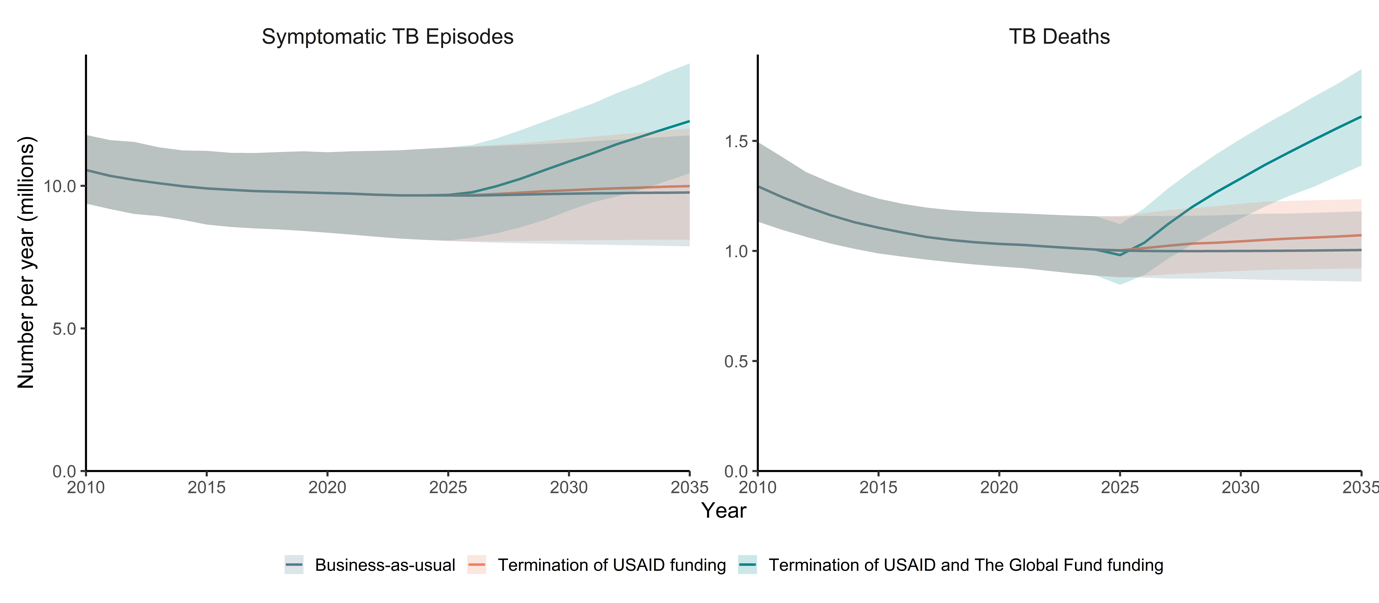

**Supplemental Figure 2**: Excess symptomatic tuberculosis (TB) episodes and TB deaths (2025–2035) projected to result from termination in funding to national TB programmes from the United States Agency for International Development (USAID) and from termination in funding to national TB programmes from USAID and The Global Fund

**Supplemental Table 2**: Excess symptomatic tuberculosis (TB) episodes and TB deaths (2025–2035) projected to result from termination in funding to national TB programmes from The Global Fund to Fight AIDS, Tuberculosis and Malaria, by donor

| **Donor** | **Excess symptomatic tuberculosis episodes** | | **Excess tuberculosis deaths** | |
| --- | --- | --- | --- | --- |
|  | **median (95% UI)** | **% (95% UI)** | **median (95% UI)** | **% (95% UI)** |
| All donors | 11 789 100  (8 669 000, 15 119 900) | 10·8%  (7·5, 15·7) | 2 974 800  (2 030 800, 3 976 300) | 26·1%  (17·7, 36·6) |
| United States | 3 093 200 (2 320 500, 3 979 400) | 2·9%  (2·0, 3·9) | 762 900 (550 700, 949 900) | 6·6%  (4·9, 8·4) |
| France | 723 700 (546, 916 100) | 0·7%  (0·5, 0·9) | 175 300 (128 200, 216 600) | 1·5%  (1·1, 1·9) |
| Germany | 537 900 (406 000, 680 700) | 0·5%  (0·4, 0·7) | 130 400 (95 400, 161 100) | 1·1%  (0·8, 1·4) |
| United Kingdom | 530 400  (400 400, 671 200) | 0·5%  (0·4, 0·7) | 128 600 (94 100, 158 800) | 1·1%  (0·8, 1·4) |
| Japan | 485 700 (366 700, 614 600) | 0·4%  (0·3, 0·6) | 117 700 (86 200, 145 400) | 1·0%  (0·8, 1·3) |
| Canada | 404 800 (305 600, 512 200) | 0·4%  (0·3, 0·5) | 98 200 (71 900, 121 200) | 0·9%  (0·6, 1·1) |
| European Commission | 316 100 (238 700, 399 900) | 0·3%  (0·2, 0·4) | 76 700 (56 100, 94 700) | 0·7%  (0·5, 0·8) |
| Sweden | 120 600 (91 000, 152 400) | 0·1%  (0·1, 0·1) | 29 300 (21 400, 36 100) | 0·3%  (0·2, 0·3) |
| Norway | 84 700 (64 000, 107 100) | 0·1%  (0·1, 0·1) | 20 600 (15 100, 25 400) | 0·2%  (0·1, 0·2) |
| Italy | 80 600 (60 900, 101 900) | 0·1%  (0·1, 0·1) | 19 600 (14 300, 24 100) | 0·2%  (0·1, 0·2) |
| Netherlands | 78 400 (59 200, 99 100) | 0·1%  (0·1, 0·1) | 19 000 (13 900, 23 500) | 0·2%  (0·1, 0·2) |
| Australia | 77 900 (58 800, 98 400) | 0·1%  (0·1, 0·1) | 18 900  (13 800, 23 300) | 0·2%  (0·1, 0·2) |
